# Effects of a Telehealth-Delivered Mediterranean Diet Intervention in Adults with Rheumatoid Arthritis (MEDRA): A Randomised Controlled Trial

**DOI:** 10.1101/2023.02.21.23286250

**Authors:** Tala Raad, Elena George, Anne Griffin, Louise Larkin, Alexander Fraser, Norelee Kennedy, Audrey Tierney

**Affiliations:** Discipline of Dietetics, School of Allied Health, Faculty of Education and Health Sciences and Health Implementation Science and Technology Cluster, Health Research Institute, University of Limerick, V94 T9PX, Ireland; Institute for Physical Activity and Nutrition (IPAN), School of Exercise and Nutrition Sciences, Deakin University, Geelong, Victoria, 3220, Australia; Discipline of Physiotherapy, School of Allied Health, Faculty of Education and Health Sciences, Implementation Science and Technology Centre, Health Research Institute, University of Limerick, V94 T9PX, Ireland; Department of Rheumatology, University Hospital Limerick, Limerick, V94 T9PX, Ireland; Graduate Entry Medical School, Faculty of Education and Health Sciences, University of Limerick, Limerick, V94 T9PX, Ireland; School of Allied Health, Human Services and Sport, Faculty of Science and Engineering, La Trobe University, Melbourne, Vic, 3086, Australia

**Keywords:** Mediterranean diet, rheumatoid arthritis, physical function, quality of life, telehealth, Ireland

## Abstract

**Objective:** To compare the effects a Mediterranean diet (MedDiet) versus the Irish Healthy Eating Guidelines (HEG) on physical function and quality of life in adults with rheumatoid arthritis (RA) in Ireland.

**Methods:** Forty-four adults with RA were randomised (1:1) to the MedDiet or HEG for 12 weeks. The intervention included three video teleconsultations and two follow-up telephone calls facilitated by a Registered Dietitian (RD). Changes in physical function by Health Assessment Questionnaire-Disability Index (HAQ-DI) and quality of life by Rheumatoid Arthritis Quality of Life Questionnaire (RAQoL) were the primary outcomes measured. Secondary outcomes included changes in dietary adherence, physical activity by Yale Physical Activity survey (YPAS), patient-perceived pain and general health, and anthropometric measures. All measurements were administered at baseline and repeated at 6 and 12 weeks.

**Results:** Forty participants completed the study. Participants were primarily females (87.5%), mean age was 47.5 ± 10.9 years. At the end of the intervention, participants in the MedDiet group reported significantly better physical function (p=0.006) and quality of life (p=0.037) compared to HEG group. From baseline to 12 weeks, physical function significantly improved in both die tgroups, MedDiet (0.9 ± 0.5 to 0.5 ± 0.4 units, p <0.001) and HEG (1.4 ± 0.7 to 1.0 ± 0.6 units, p<0.001). Quality of life also significantly improved in the MedDiet (10.1 ±7.5 to 4.0 ± 4.7 units, p<0.001) and HEG group (11.25 ±7.2 to 7.9 ±6.4 units, p=0.048). Physical activity improved significantly in the MedDiet (56.7 ±28.6 to 70.6±33.5 points, p=0.01) but not within the HEG group despite similar recommendations.

**Conclusion:** Adhering to the MedDiet and Irish Healthy Eating Guidelines resulted in improvements in RA patient-reported outcomes. The changes observed in both diet groups are likely due to the improvement in overall diet quality irrespective of dietary prescription.

**Trial registration number:** NCT04262505

## Introduction

Rheumatoid Arthritis (RA) is the most common type of autoimmune arthritis affecting between 0.5 -1% of the adult population worldwide [1]. At present, it is thought that up to twenty three million people worldwide are living with RA [2]. In Ireland, it is estimated that more than 40,000 people are currently living with the condition [3]. RA can affect people of any age including children, but it is most common between 40 and 60 years of age [4]. Females are affected by the condition nearly three times more than men [4]. RA is characterised by progressive joint inflammation, damage, and disability, all of which severely impact the person’s quality of life [5]. RA manifests through various symptoms including pain, morning stiffness, fatigue, as well as limited mobility, all of which can affect mental health and wellbeing [4, 6]. People with RA experience progressive worsening in their physical function, limiting their ability to carry out daily life activities and affecting their quality of life [7].

People with RA have 50% higher risk of developing cardiovascular disease (CVD) compared to the general population, and CVD is considered one of the leading causes of death for adults with RA [8]. As there is no cure for RA, the overall goal of treatment is to relieve pain, decrease inflammation and prevent joint damage in order to preserve function and quality of life [9]. Treatment options for RA commonly include pharmacological strategies, physical and occupational therapies, and surgical interventions [10]. However, many people report many side effects or inadequate response to pharmacologic therapies [11]. Therefore, a wholistic approach that includes both pharmacologic and nonpharmacologic therapies must be considered for optimal care. Despite the lack of consensus regarding nonpharmacologic therapies, people with RA continue to express interest in adopting self-management strategies and lifestyle modifications. Recent efforts have been made to improve lifestyle behaviours in people with RA. In 2021, the European League Against Rheumatism (EULAR) formulated principles and recommendations regarding lifestyle behaviours to prevent disease progression in people with rheumatic and musculoskeletal conditions. These principles emphasise the importance of eating a healthy balanced diet, maintaining a healthy weight and exercising on a regular basis [12].

People living with RA are often interested in modifying their diet as a way to help them manage their symptoms and cope with their condition [13]. It has been reported that 33–75% of people with RA believe that diet affects the severity of their symptoms and approximately 50% have tried to change their diet in an attempt to alleviate their symptoms [14]. In many cases, these dietary changes are made without any input from a healthcare professional [15]. The role of diet in the progression, and alleviation of RA-related symptoms has been investigated in a number of studies. Several dietary patterns including vegetarian, vegan, elimination, elemental, ketogenic, and the Mediterranean Diet (MedDiet) have been examined [13, 16, 17]. Whilst some studies have demonstrated positive effects, other studies have shown no change [17]. Therefore, to date, no one diet has been determined as being superior or most effective in the management of RA and diet remains excluded from the clinical practice guidelines for RA treatment.

The anti-inflammatory and antioxidant potential of a MedDiet is thought to delay the onset and halt the progression of inflammation in chronic inflammatory conditions [18]. The dietary pattern has also been associated with the prevention and management of CVD [19]. Given that RA has an underlying inflammatory pathogenesis and coexists with CVD, and to address the unmet need of an optimal dietary prescription for people with RA, the French Society for Rheumatology (SFR) published the first set of dietary recommendations for chronic inflammatory rheumatic diseases. The recommendations support the adoption of a MedDiet to help relieve RA-related symptoms and improve outcomes [20]. In line with this, a systematic review and meta-analysis, published in 2021, assessing the effects of diets with anti-inflammatory basis in RA, reported that a MedDiet seemed to have more beneficial effects on pain compared to vegan and vegetarian diets [21]. This is further supported by a recent systematic review that investigated the effect of a MedDiet on RA-related outcomes and demonstrated several health benefits including improvements in pain and physical function [22].

In Ireland, the current healthy eating guidelines (HEG) encompass the principles of a low-fat, high fibre diet with emphasis on optimal portion sizes to promote and maintain health. There is limited evidence on the effects of such a diet in RA [23]. However, in a study that examined the effects of a low-fat, vegan diet in RA, participants who followed the diet experienced significant improvements in several RA parameters including pain, morning stiffness and inflammatory biomarkers, however, the sustainability of these improvements was not reported [24].

The large body of literature surrounding the anti-inflammatory benefits of a MedDiet has created a particular interest in the use of this dietary pattern for people with RA. However, to date, limited trials have investigated the MedDiet in RA. These studies compared the effects of a MedDiet to either a typical Western diet [25], fasting [26], or written resources on healthy eating without any dietetic supervision [27]. The studies were heterogeneous in terms of design and clinical outcomes evaluated with some methodological limitations. When compared with the control group, the studies indicated that participants following the MedDiet experienced improvements in several parameters including pain, physical function, morning stiffness, quality of life and inflammatory biomarkers. Considering the favourable, albeit limited data in RA, well-conducted trials investigating the potential of a MedDiet to improve RA outcomes are warranted. The primary aim of the MEDRA study is to assess the effectiveness of a dietetic led intervention comparing the MedDiet to the Irish Healthy Eating Guidelines, (given that this is recommended by the Irish Health Authorities as an evidence-based approach to healthy eating and is likely more familiar to an Irish population), on physical function and quality of life in adults with RA in Ireland.

## Methods

### Study design

The MEDRA study was an open-label, telehealth-delivered, randomised controlled trial with two parallel intervention arms. In response to the pandemic and following the Irish government’s announcement of a national lockdown, we sought to use telehealth methods to deliver the proposed intervention [28]. The study was designed in accordance with and adheres to the Consolidated Standards of Reporting Trials (CONSORT) statement [29]. All procedures involving patients were approved by the Education and Health Sciences Research Ethics Committee at the University of Limerick (2020_09_05_EHS) and by the Health Service Executive Mid-Western Regional Hospital Research Ethics Committee (REC Ref 103/19). The 12-week dietary intervention included data collection and consultations at baseline, 6 and 12 weeks. The published protocol details the study design and methods [30]. All participants gave their written, informed consent to take part in the study. The study was registered prospectively at ClinicalTrials.gov on April 2nd, 2020 (NCT04262505). Patients and the public were not involved in the design, conduct, reporting or dissemination plans of this present study.

### Participants

Between November 2020 and February 2021, participants were recruited through social media platforms of the patient organisation ‘Arthritis Ireland’ and through outpatient rheumatology clinics at University Hospital Limerick (UHL) via poster advertisement. Eligible participants were adults (≥18 years old) with a RA diagnosis fulfilling the 2010 American College of Rheumatology/ European League against Rheumatism (ACR/EULAR) RA classification criteria [31]; and had access to the internet or a mobile device capable of receiving text messages and telephone calls. Exclusion criteria were commencement of nutritional supplements or a new dietary regime in the month prior to study enrolment, pregnancy, breastfeeding, and non-English speaking.

Participants who expressed interest in the study were screened for initial eligibility via a telephone call by a CORU (Irish regulating Health and Social Care Professional) Registered Dietitian (RD) (TR). Eligible participants were provided with a detailed study information sheet and consent form via post or email. A week later, potential participants were contacted by the RD via email and subsequently scheduled to attend a baseline video teleconsultation upon providing written consent. All participants continued to receive standard medical care provided at their respective hospital or primary care settings; however, participants were advised not to engage in any weight-loss or other lifestyle interventions that might affect the study.

### Randomisation and blinding

A computer-generated blocked randomisation list allocated participants 1:1 to either the MedDiet or the HEG group. Randomisation was conducted by a statistician who is not involved in the study. Due the nature of dietary intervention studies, both, participants, and the RD could not be blinded. All assessments and analyses were performed by the same RD not blinded to allocation. Therefore, to minimize the risk of bias, all data were fully pseudomised before analysis.

### Interventions

Both dietary interventions comprised personalised dietary advice and nutrition counselling including nutrition education and goal setting from the RD to help improve dietary quality and achieve adherence to the prescribed diet. While both dietary interventions were standardised to the overarching principles of the assigned diet, the RD adopted a tailored approach with regards to assisting individuals in achieving lasting dietary behavior change based on individuals’ likes and dislikes. All participants were made aware that this was not a weight loss intervention and were advised to remain weight stable through regular at-home weight monitoring. Participants in both diet groups were encouraged to engage in regular physical activity (at least 30 minutes a day of moderate intensity activity, five days a week). A log was kept by the RD whereby participants dietary preferences and food allergies were recorded.

### Teleconsultation schedule

The RD delivering the intervention (TR) informed all study participants of their group allocation during the baseline video teleconsultation. Both diet groups received three video teleconsultations (baseline, 6, and 12 weeks) and two follow-up phone calls at weeks 3 and 9. All calls were scheduled on weekdays at a time of the participants choosing. The video teleconsultations lasted approximately 40 minutes while the follow-up phone calls took between 15 and 20 minutes.

### MedDiet

The MedDiet intervention was designed based on the principles of the traditional Cretan MedDiet [32]. Based on to the MedDiet food pyramid, participants were advised to consume 60–80 ml of EVOO, at least two servings of vegetables and three servings of fruits daily. Participants were also instructed to include fish at least 3 times per week and a handful of raw unsalted nuts every other day. Participants in the MedDiet group were provided with a MedDiet resource handbook which was developed based on the MedDiet guide by George et al. [33]. The handbook included a 2-wk meal plan modeling the MedDiet and its key dietary components and a combination of traditional and adapted recipes considered to be suitable in an Irish setting. The handbook included a food pyramid, a shopping list and general advice on how to follow and adhere to the main principles of the MedDiet. The handbook was adapted from resources that have been successfully used in previous MedDiet intervention studies in non MedDiet countries that the authors have been involved in [33].

## HEG

Participants in the HEG group were instructed to follow the Irish Healthy Eating Guidelines published by the Department of Health in December 2016. In this intervention group, participants were requested to consume 5-7 servings/ of fruits and vegetables, 3-5 servings of wholegrain cereals, 3 servings of low-fat dairy, 2 servings of lean meat. Participants were asked to limit the intake of high fat, sugar, salt food and drinks to once or twice/wk and to use fats, spreads, and oil in very small amounts. All participants were supplied with a 101 Square Meals recipe book [34]. Sample daily meal plans were readily available online for participants on the Healthy Ireland website (HSE, http://www.hse.ie).

### Dietary assessment

#### 3-day food diary

Dietary data was collected using three-day food diaries to assess adherence to dietary prescription. Food diaries included two weekdays and one weekend day and were collected at baseline, 6 and 12 weeks. Participants were requested to complete the food diaries using the LIBRO mobile application^®^. The three-day food diaries were then imported into and nutrition analysis was completed by the RD using the Nutritics Software^®^.

#### Dietary adherence scores

The Mediterranean Diet Adherence Score (MEDAS) is a 14-point short screener developed and validated by researchers for the PREDIMED trial, a study which examined the effects of a MedDiet in individuals with CVD [35]. The checklist includes the key principles of the MedDiet. The score is intended to assess adherence to the MedDiet where scores range from 0 to 14 points. Adherence is categorised into three categories: low (scores were between 0– 5), moderate (scores 6–9) and high (scores 10–14) [36]. All participants in the MedDiet group completed the checklist at baseline, 6 and 12 weeks.

In the HEG group, an 11-item healthy eating checklist specifically developed by the authors was used to crosscheck against and assess adherence to the Irish Healthy Eating Guidelines. Since there is an overlap in dietary prescription between the MedDiet and the HEG in terms of certain food groups (vegetables, fruits, wholegrains etc..), adherence to the MedDiet was also assessed in the HEG group using the 14-item MEDAS.

### Primary and Secondary Outcomes

The primary endpoints were changes in physical function and quality of life at week 12. Both measures were assessed using self-administered validated questionnaires, Health Assessment Questionnaire -Disability Index (HAQ-DI) [37] and Rheumatoid Arthritis Quality of life (RAQoL) questionnaire [38], respectfully, with lower scores for both questionnaires indicating better physical function and quality of life. Secondary endpoints included changes observed in patient-perceived pain and general health assessed using the HAQ-DI. Physical activity was measured using the validated Yale Physical Activity Survey (YPAS) [39]. All assessments were carried out at baseline, 6 and 12 weeks.

### Statistical analysis

The data set was analysed with an intent-to-treat (ITT) principle and used the last observation carried forward method for missing data. Descriptive statistics were completed and reported as mean ± SD. Normality of data was assessed using the Shapiro–Wilk test with a non-significant result (p > 0.05) indicating normality. Categorical variables were expressed as frequencies and percentages. The level of statistical significance was set at p < 0.05. Group differences were calculated using the chi-square test for categorical variables. The independent samples t-test was used to compare the mean scores between two groups of normally distributed continuous variables, and the Mann-Whitney U test was employed as the non-parametric alternative. Paired samples t-test was used to compare the mean scores within groups between baseline and end of intervention assessment. The Wilcoxon Signed Rank Test was used as the non-parametric alternative. A general linear model, i.e., repeated-measures ANOVA (analysis of variance), was used to examine the between-group differences of mean values at each time point of measurement, the within-group changes (time effect) from baseline to follow up in each intervention group, and the differences in the changes from baseline to follow up between the two intervention groups (treatment × time interaction effect). All statistical analysis was performed using the Statistical Package for the Social Sciences SPSS statistical software for Windows (IBM, version 27).

### Power calculation

The study was designed to provide 80% power to detect a difference of 0.68 units in HAQ-DI score (clinically relevant) [40] from baseline, assuming a two-sample Student’s t-test and α = 0.05 significance level. The calculated sample size was 40 participants in total. Adjusting for a potential 10% dropout, the sample size was determined to be 44 participants.

## Results

Of the 57 patients screened for eligibility, 44 participants were recruited and randomly assigned to receive either the MedDiet (n=22) or HEG (n=22). Of the 44 participants enrolled, 40 completed the study (Figure 1). Reasons for exclusion and withdrawals for all screened patients are shown in Figure 1.

**Figure 1.**
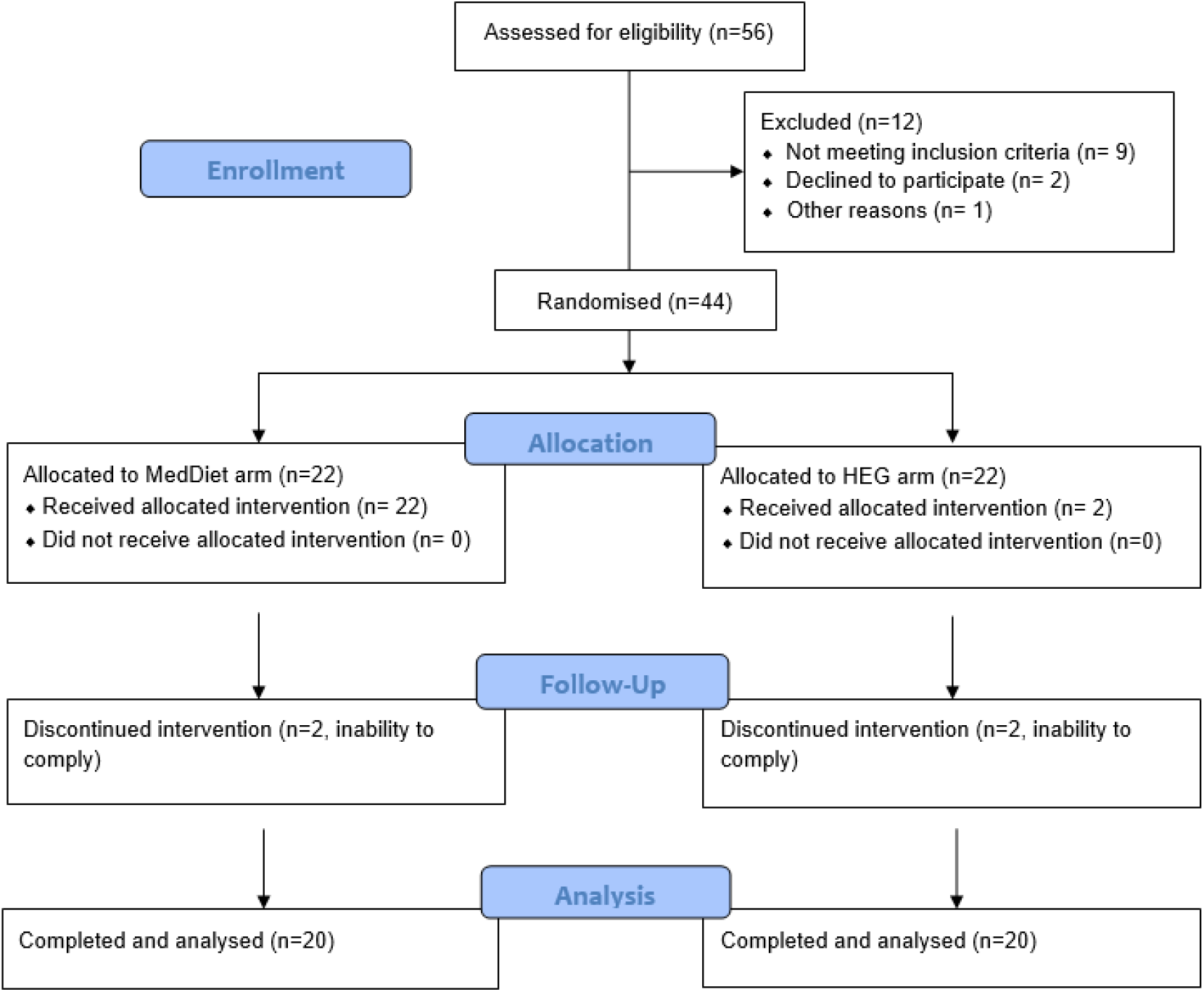
MEDRA study participants flow diagram.

### Baseline demographics and characteristics

Table 1 presents the characteristics of study participants in terms of their socio-demographics, anthropometric and patient reported outcomes in the total sample (n=40) and by diet group. Study participants had a mean ± SD age of 47.5 ± 10.9 years old (range, 23-70 years old) with the cohort being predominantly female (87.5%), reflective of a typical RA population [41]. Most participants recruited (92.5%) were born in Ireland and had completed some form of tertiary education (85%). Almost all participants (97.5%) were on disease modifying anti-rheumatic drugs (DMARDs) and/or biologic agent. At baseline, there were no differences between the two diet groups in terms demographic characteristics, anthropometric or functional variables except for the HAQ-DI score which was significantly lower in the MedDiet group indicating better baseline physical function in the MedDiet group compared to HEG group (0.9 ± 0.5 vs. 1.4 ±0.7, p<0.001) and therefore was adjusted in further analysis (Table 1).

**Table 1.**
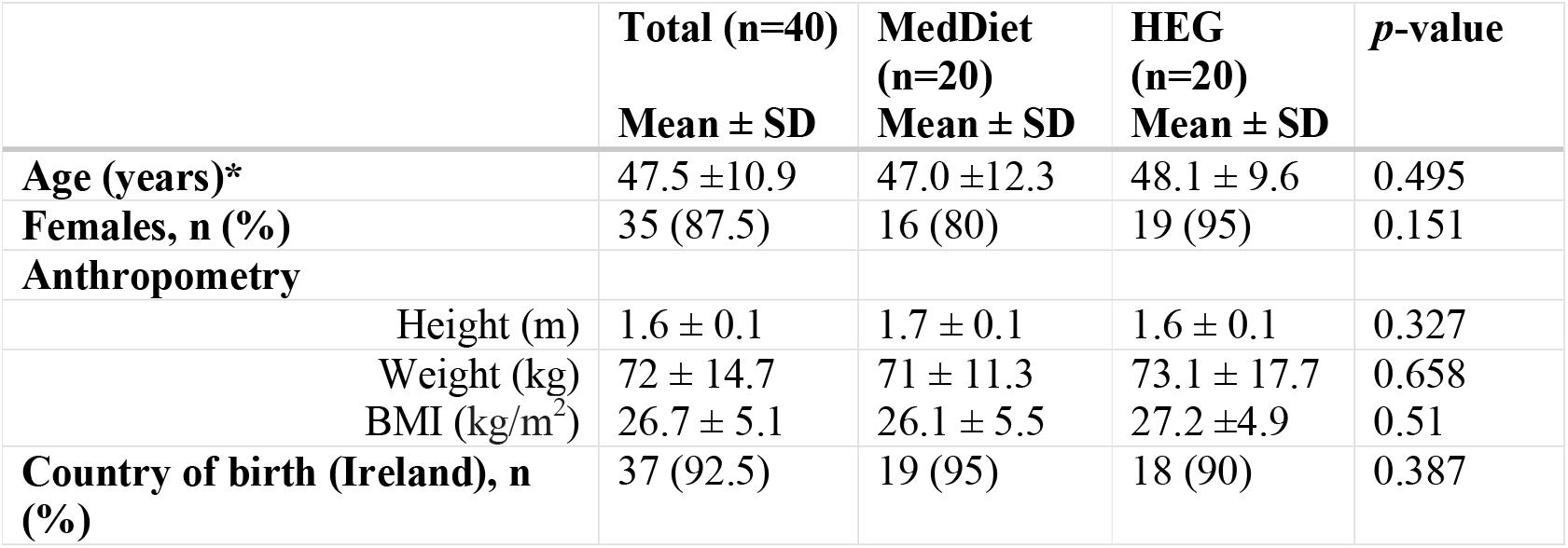
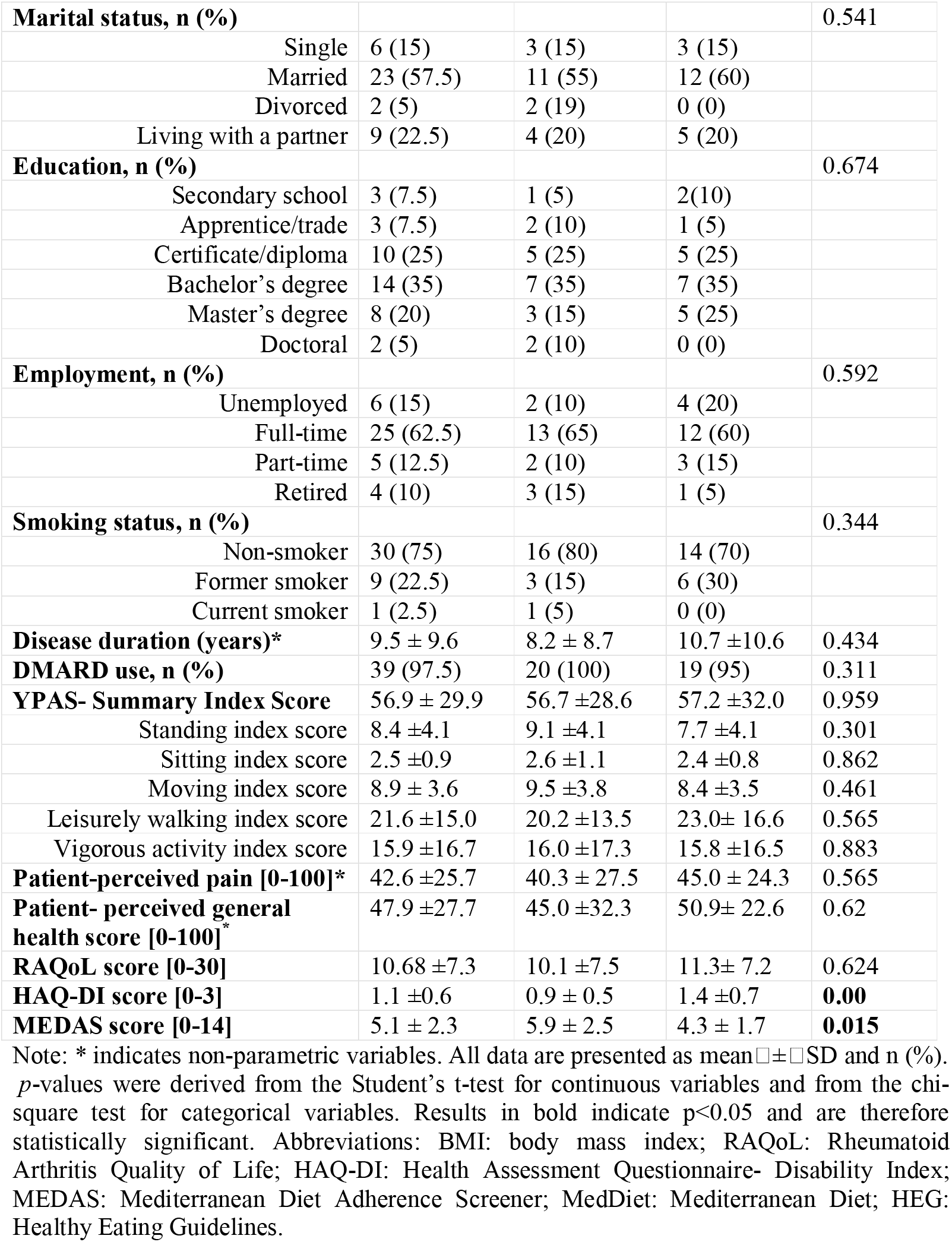
Baseline characteristics of study participants

### Effect of dietary interventions on physical function and quality of life

Taking into account the baseline differences in HAQ-DI scores between the two groups at baseline, change levels were assessed, wherein there was no significant difference between the groups (p=0.586). The change in RAQoL score from baseline to 12-wk was also non-significant between the two groups (p=0.165) (Table 4). However, at 12 weeks, participants allocated to the MedDiet group reported significantly lower HAQ-DI and RAQoL scores compared to participants in the HEG group; HAQ-DI (p=0.006) and RAQoL (p=0.037) indicating better physical function and quality of life

### Effect of dietary interventions on physical activity and patient-perceived pain and general health

No significant between-group differences were observed for reported pain and health status at the end of the intervention (Table 4). However, when assessed within group there was a significant improvement in pain and health status from baseline to 12 weeks, pain: (MedDiet: 40.3 ± 27.5 to 17.4 ± 22.2, p<0.001; HEG: 45.0 ± 24.3 to 30.3 ± 30.1, p=0.028), health status: (MedDiet: 45.0 ±32.3 to 29.0 ±34.9, p=0.001; HEG: 50.9 ±22.6 to 28.7 ±24.1, p=0.002). Physical activity levels were significantly higher among participants in the MedDiet group compared to the HEG group at the end of the intervention (p<0.001). From baseline to post-intervention, there was a significantly increase in physical activity levels in the MedDiet (56.7 ± 28.6 to 70.6 ± 33.5 points, p=0.01) and a non-significant decrease in the HEG group (57.2 ± 32.0 to 54.3 ± 23.8 points, p=0.618).

### Effect of dietary interventions on anthropometric measures

The effect of the two dietary interventions on weight and BMI is presented in Table 4. There were no significant differences for post-intervention values for weight and BMI between the two diet groups. There were also no significant differences noted for the changes observed from baseline and 12 weeks between the two diet groups (Table 4). Although this was not a weight loss study, during the intervention period, there was a small and significant reduction in body weight among both groups when within group changes were assessed. Weight loss in participants who followed the MedDiet was -0.6 kg (71.0 ±11.3 kg to 70.4 ±11.5 kg, p<0.001) and those who followed the HEG was -0.9 kg (73.1 ±17.7 kg to 72.2 ±17.3 kg, p=0.004) from baseline to 12-wk. Correspondingly, there was a significant reduction in BMI (kg/m^2^) in the MedDiet (26.1 ±5.5 to 25.9±5.5 Kg/m^2^, p=0.001) and HEG (27.2±4.9 to 26.9±4.7 Kg/m^2^, p=0.002) group over the 12-wk intervention period.

### Baseline adherence to the assigned diet

Overall, participants in the MedDiet group had low baseline adherence to a MedDiet dietary pattern with a mean MEDAS score of 5.9 ± 2.5 out of 14. The HEG group had a mean adherence score of 6.3 ± 1.7 out of 11 to Irish Healthy Eating Guidelines. When adherence to the MedDiet was assessed among participants in the HEG group at baseline using the MEDAS, participants also had low baseline adherence to the MedDiet dietary pattern with a mean MEDAS score of 4.3 ± 1.7 out of 14. Accordingly, the pooled cohort of participants had low baseline adherence to the MedDiet pattern, with a mean MEDAS score of 5.1 ± 2.3 out of 14 (Table 1).

### Adherence to the dietary intervention from baseline to 12 weeks

The increase in the MEDAS score in the MedDiet group was significant across all three time points (Figure 2). The increase in the HEG score in the HEG group was significant between baseline and 6 weeks (p<0.001) and between baseline and 12 weeks (p<0.001) (Figure 3). Thus, indicating that all participants adhered to the prescribed advice within each respective diet group met their assigned dietary prescriptions, improving their diet quality.

**Figure 2.**
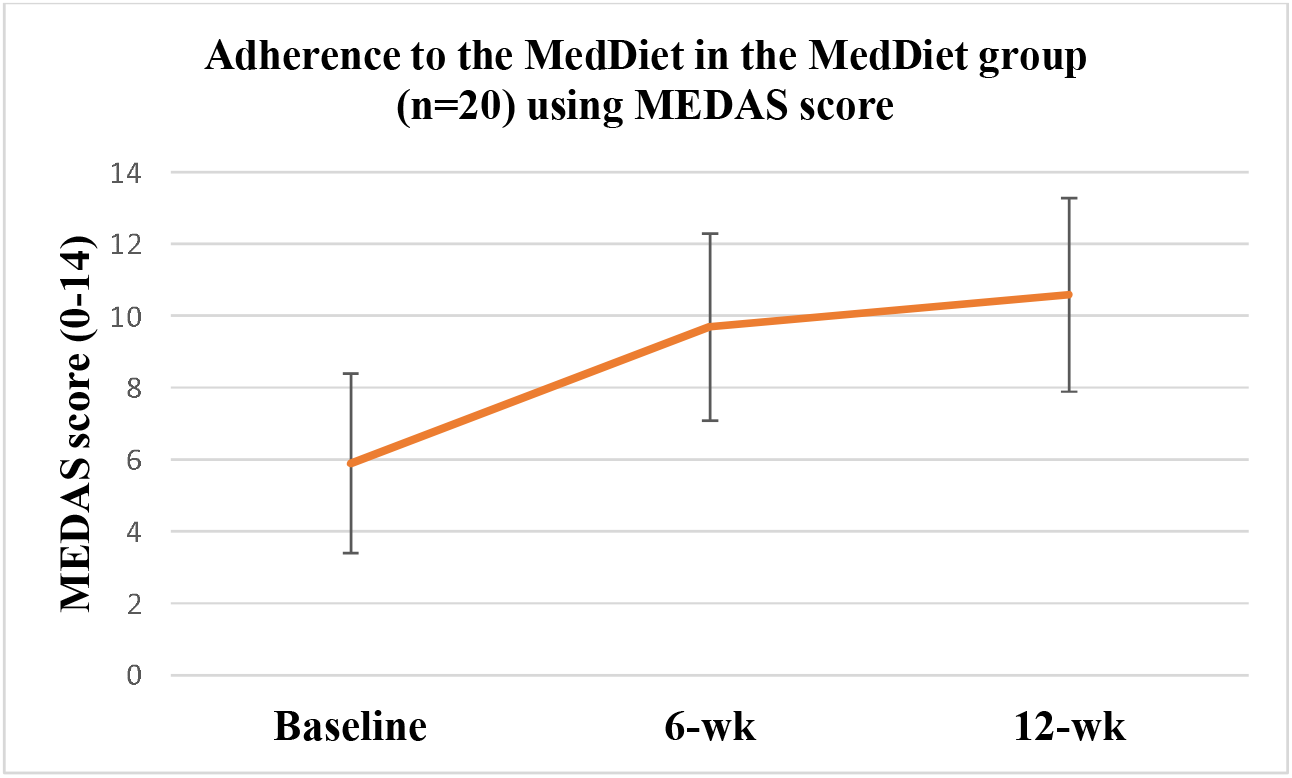
Adherence to the MedDiet principles in the MedDiet group using 14-item MEDAS.

**Figure 3.**
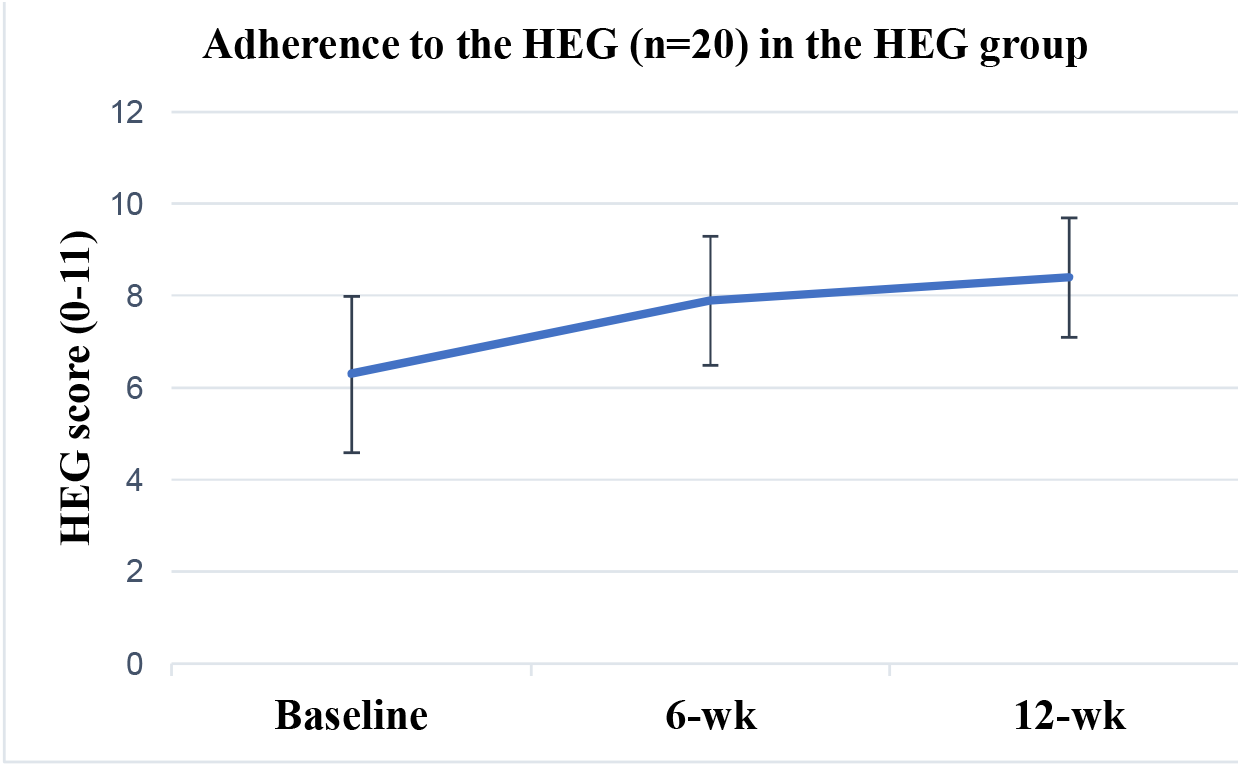
Adherence to the HEG in the HEG group using 11-item Healthy Eating checklist.

To assess overall diet quality and adherence to MedDiet principles total MEDAS scores across all time points in both groups are presented in Figure 2. At 12-wk, both diet groups had significantly higher MEDAS scores compared to baseline, MedDiet (5.9 ± 2.5 to 10.6 ± 2.7, p<0.001) and HEG (4.3 ± 1.7 to 6.9 ±1.5, p<0.001). The change from baseline to post-intervention was significantly higher in the MedDiet group compared with the HEG group (4.7 ± 3.2 vs. 2.6 ± 1.7, p=0.017), respectfully (See Table 2).

**Table 2.** Adherence to the MedDiet at pre- and post-intervention in the pooled cohort and by diet arm using the MEDAS.

|  | Baseline | 12 wk | Within group p-value | Mean Change pre-post (95% CI) | Mean change between groups p-value |
| --- | --- | --- | --- | --- | --- |
| **Cohort**<br>**(n=40)** | 5.1 ± 2.3 | 8.7 ± 2.9 |  | 3.6 ± 2.7 | **0.017** |
| **MedDiet**<br>**(n=20)** | 5.9 ± 2.5 | 10.6 ± 2.7 | **<0.001** | 4.7 ± 3.2 |  |
| **HEG (n=20)** | 4.3 ± 1.7 | 6.9 ± 1.5 | **<0.001** | 2.6 ± 1.7 |  |

Abbreviations: MEDAS: Mediterranean diet adherence screener; MedDiet: Mediterranean Diet; HEG: Healthy Eating Guidelines, CI: confidence interval. All data are presented as mean ± SD. Results in bold indicate p<0.05 and are therefore statistically significant.

**Table 3.** Anthropometric and patient-reported outcomes across intervention time points.

|  | MedDiet (n=20) |  |  |  |  | HEG (n=20) |  |  |  |  | Mean change between groups p-value |
| --- | --- | --- | --- | --- | --- | --- | --- | --- | --- | --- | --- |
|  | Baseline | 6 wk | 12 wk | Mean Change pre-post (95% CI) | Within group p-value | Baseline | 6 wk | 12 wk | Mean Change pre-post (95% CI) | Within group p-value |  |
| **Wt (kg)** | 71.0 ±11.3 | 70.2 ±11.6 | 70.4 ±11.5 | -0.6 ±0.7 | <0.001 | 73.1 ±17.7 | 72.0 ±17.3 | 72.2 ±17.3 | -0.9 ±1.2 | <0.001 | 0.472 |
| **BMI (kg/m<sup>2</sup>)** | 26.1 ±5.5 | 25.8 ±5.5 | 25.9 ± 5.5 | -0.2 ±0.3 | <0.001 | 27.2±4.9 | 26.8 ±4.7 | 26.9±4.7 | -0.3±0.4 | <0.001 | 0.461 |
| **Physical function HAQ-DI score (0-3)** | 0.9 ±0.5 | 0.7±0.4 | 0.5 ±0.4 | -0.3±0.3 | <0.001 | 1.4±0.7<sup>a</sup> | 1.2±0.7 | 1.0±0.6 | -0.4±0.4 | <0.001 | 0.586 |
| **Quality of life RAQoL score (0-30)** | 10.1 ±7.5 | 6.9±5.9 | 4.0 ± 4.7 | -6.1±6.1 | <0.001 | 11.25 ±7.2 | 9.35±6.5 | 7.9 ±6.4 | -3.4±5.9 | 0.048 | 0.165 |
| **HAQ-DI pain (0-100) \*** | 40.3±27.5 | 22.1±28.4 | 17.4±22.2 | -22.9±21.4 | <0.001 | 45.0 ±24.3 | 29.7±27.9 | 30.3 ± 30.1 | -14.7±33.1 | 0.028 | 0.363 |
| **HAQ-DI**<br>**General**<br>**health (0-**<br>**100) \*** | 45.0<br>±32.3 | 27.8±32.9 | 29.0 ±34.9 | -16.0±33.9 | **<0.001** | 50.9 ±22.6<sup>a</sup> | 35.3±31.4 | 28.7 ±24.1 | -22.2±24.8 | **0.002** | 0.513 |
| **Physical**<br>**activity**<br>**YPAS**<br>**score**<br>**(0-142)** | 56.7<br>±28.6 | 66.9±27.3 | 70.6±33.5 | 13.9±21.5 | **0.009** | 57.2 ±32.0 | 53.0±21.3 | 54.3 ±23.8 | -2.9±25.6 | 0.721 | **0.031** |

Note: \* indicates non-parametric data, statistical analysis conducted using non-parametric tests. Within-group values were compared with paired t test. Differences between groups were analysed by Student's t test for independent samples. Abbreviations: MedDiet: Mediterranean Diet, HEG: Healthy Eating Guidelines Wt: weight; BMI: Body mass index; HAQ-DI: Health Assessment Questionnaire-Disability Index; RAQoL: Rheumatoid Arthritis Quality of Life; YPAS: Yale Physical Activity Survey. Results in bold indicate p<0.05. Lower HAQ-DI score indicates better physical function, lower pain score indicates less pain, lower health status score indicates better health status, lower RAQoL score indicates better quality of life.

## Discussion

The MEDRA study was designed to investigate the effects of a MedDiet compared to the Irish Healthy Eating Guidelines on physical function and quality of life in a cohort of adults with RA. The study demonstrated that significant improvements in patient-reported outcome measures namely physical function, quality of life, patient-perceived pain and general health can be achieved in adults with RA through the improvement of diet quality via both, a MedDiet and the Irish HEG. The observed improvements in these parameters suggest that dietary management is effective in improving RA-related outcomes and therefore should be considered alongside other, proven management strategies including medical treatments and physiotherapy.

Results from the MEDRA study demonstrated that adherence to the MedDiet intervention, delivered by a RD, resulted in improvements in physical function, quality of life, patient-perceived pain and general health. These findings are in keeping with previous MedDiet intervention studies in RA. Sköldstam et al. in (2003) reported significant improvements in physical function, patient-reported quality of life and disease activity score (DAS-28) in participants who followed a MedDiet for 12 weeks while participants who followed a typical Western diet did not show any improvements [25]. McKellar et al. (2007) conducted a pilot study among female RA patients and found that participants assigned to the MedDiet for three months demonstrated significant improvements in physical function and patient-reported pain [27]. Together, these studies highlight potentially beneficial effects from the Mediterranean dietary pattern for the management of RA.

The beneficial effects of the MedDiet have been attributed to its various components rich in antioxidants and anti-inflammatory properties [42]. It has been proposed that the MedDiet can alter the gut microbiome, thus modulating the inflammatory response in RA [43]. Dietary components of MedDiet can affect disease activity in RA through direct or indirect interactions with the immune system. Epidemiological studies suggested that anti-inflammatory properties of flavonoids and carotenoids, widely included in MedDiet, inhibit both isoforms of inducible nitric oxide synthase (iNOS) and of cyclooxygenase 2 (COX-2), which are which are responsible for producing inflammatory mediators [44]. Another key element of the MedDiet is the central aspect of extra-virgin olive oil, which due to its high mono-unsaturated fatty acid (MUFA) content, exhibits a capability to reduce oxidative stress, and is associated with a reduction of thromboxane which induces platelet aggregation and arterial constriction [45]. In accordance with this, a study including 208 consecutive patients with RA, reported that the DAS-28 was significantly correlated with the ratio of consumed monounsaturated to saturated fatty acid (MUFA/SFA) [46]. Moreover, the MedDiet emphasises moderate intake of fish, especially those rich in omega-3 fatty acids such as salmon, tuna, etc.. The omega-3 fatty acids inhibit arachidonic acid metabolism, which results in the synthesis of less thrombogenic and less inflammatory eicosanoids such as TNF-alpha and interleukin-1 beta (IL-1 beta) responsible for to tissue destruction and pain in RA [47].

A major finding from the present study was that an Irish cohort of adults with RA achieved high adherence to a MedDiet after a 12-wk RD led intervention. At baseline, participants in both diet groups had low adherence to a MedDiet. Despite the increased familiarity of the MedDiet through its promotion as the most widely evidence-based diet for health, this finding was expected given that the Irish Healthy Eating Guidelines, familiar to most, are based around a low-fat, high fibre dietary pattern. When group adherence to the MedDiet was assessed at 12-wks, improvements indicating higher levels of adherence were achieved in both diet groups. Changes in MedDiet adherence using the MEDAS score, showed a mean increase by 3.6 out of 14 points in the overall group (MedDiet: 4.7 points; HEG: 2.6 points).

This significant improvement in adherence to the MedDiet principles in the MEDRA cohort is an important finding and is indeed encouraging as it indicates that it is possible to adhere to principles of a MedDiet in non-Mediterranean populations. While some components of the MedDiet overlap with other healthy dietary patterns [48], it remains unclear from previous studies whether a MedDiet as a whole is superior in RA with regards to positive outcomes being achieved or if a standard healthy diet complemented with key elements of a MedDiet would yield similar benefits. It is also important to note that whilst the MedDiet is recognised as one of the healthiest diets in the world [49], it is not the only healthy eating pattern that exists and that is backed by scientific research. Different countries have their own dietary guidelines that promote healthy diets and are similar to the MedDiet in that they promote variety, moderation and focus on whole foods. These dietary recommendations are often tailored to the population, and take into account the country’s food availability and food culture [50]. Therefore, when both diets yield similar results, it is important to consider what is more convenient and familiar to the participants.

While weight loss was not intended or anticipated for participants enrolled in this trial, both diet interventions resulted in small but significant weight reduction. In this study, the change in body weight was similar in both groups whereby the MedDiet group lost 0.6 kg and the HEG group lost 0.9 kg over the 12-wk intervention period. The increased adherence to the respective dietary prescriptions and improvement in diet quality in both diet groups may explain this reduction in body weight. These results align with results from other MedDiet intervention studies whereby, Sköldstam et al reported that participants in the MedDiet group lost 3.0 kg in weight (p<0.001) and participants in control group remained weight stable throughout the 12-wk study period [25]. In Mckellar et al.’s study, the MedDiet group lost 0.9 kg whereas the group receiving tips on healthy diet showed a 3 kg weight gain over the 6-month study period [27]. Several studies have demonstrated that participating in behavioural lifestyle interventions often lead weight loss [51]. Weight loss has commonly been reported in response to dietary intervention for patients with RA [16]. However, there is a paucity of studies assessing the effects of weight loss in people with RA. Evidence of weight loss reported in response to dietary intervention in patients with RA have seen no association between the patient’s decrease in body weight and the positive clinical results achieved [52]. Moreover, according to 2021 EULAR recommendations regarding lifestyle behaviours, weight loss is only advocated for patients who are overweight or obese [12].

Studies have shown that physical activity levels among RA patients, particularly among those above the age of 55 years, is lower than the level recommended by international guidelines for health-enhancing physical activity and is also lower than that among healthy persons [53]. In this study, although regular physical activity was emphasised in both dietary intervention groups, improvement in physical activity levels were only found among participants assigned to the MedDiet. This perhaps may be explained by the lower baseline YPAS score in the MedDiet group which provided more room for improvement compared to the HEG group. The improvements in physical activity levels that accompanied the changes in MedDiet adherence were anticipated given that participation in behavioural and lifestyle interventions tend to have a positive effect on individuals’ wellbeing, particularly improving physical activity and mental health [54-56]. Interestingly, a recent study reported that a MedDiet combined with a dynamic exercise programme could result in greater improvements in quality of life compared to MedDiet alone among adults with RA [57]. With that said, the superior effects observed in physical function and quality of life at week 12 in the MedDiet group in our study could have been supported by the increase in physical activity levels among participants assigned to this group.

The main strengths of this study are its randomised controlled trial design. In addition, the researchers included robust collection of a 3-day food diary to assess compliance to the dietary prescription which was checked and validated by a qualified RD who also provided telehealth nutritional counselling and strict follow-up throughout the intervention period. Furthermore, participants continued their habitual pharmacologic treatment, making the results more representative to the patient population. The use of medication was self-reported at baseline and any changes during the intervention period were documented. The validated 14-MEDAS scoring instrument from the large Spanish PREDIMED trial was used to measure compliance and adherence to the MedDiet. Primary outcomes were also robust including the validated, gold standard HAQ-DI for measuring physical function and the official Swedish RAQoL for quality-of-life measure.

The mode of healthcare delivery has shifted with exceptional speed due to Covid-19 pandemic. Telehealth has been applied as a solution to provide patient care during this period [58]. While telehealth-delivered clinical trials and dietary interventions conducted specifically in RA are lacking, trials conducted in the other chronic disease population have demonstrated that telehealth-delivered dietary interventions are effective at supporting behavioural change [59, 60]. Particularly, telehealth has demonstrated greater improvements in dietary behaviours and clinical outcomes when compared with usual care in people with chronic conditions [61, 62].

It is important to consider the limitations of this trial. Firstly, the sample size of the cohort, despite being powered on physical function, could limit our ability to assume the results will apply to the RA population and particularly to men with RA. The intervention period was for 12 weeks and while comparable to similar studies [25], however, is not reflective of sustainability of dietary and health related outcomes. Furthermore, in dietary interventions it is not possible to blind clinicians and participants from the dietary prescriptions. While data was de-identified, the RD who carried out the assessments also carried out the analysis, therefore, blinding was not possible.

While the MEDRA study benefits from the use of a disease-specific patient-reported outcomes, the subjective nature of patient-reported outcomes and self-reported dietary intake assessment methods presents numerous challenges to obtaining accurate information When dietary data is self-reported by participants, inconsistency, and under-reporting of food and/or beverage intake frequently occurs. Therefore, the possibility of beneficial effects in dietary intervention groups need to be substantiated in large, controlled trials with objective measures such as inflammatory biomarkers and disease activity score. Though 3-day food diaries are less prone to recall bias and poor reliability compared to other assessment methods such as food frequency questionnaires and 24-hour recalls, it is crucial to note that there is always potential for over/underreporting due to knowledge and awareness particularly in overweight and obese study participants Due to Covid-19 related lockdowns and government restrictions, we were unable to collect bloods for assessment of inflammatory markers or relevant dietary biomarkers, hence, limiting objective markers and exploration into the physiological drivers of the outcomes reported.

Given the significant impacts of RA on the physical, psychological, economic, and social aspects of the lives of people living with the condition, the management of RA must extend its focus beyond pharmacological treatment [63]. The MEDRA study has shown that improving diet quality offers potential benefits across a range of parameters, suggesting that this approach may be a useful therapeutic adjunct to traditional DMARDs. To our best knowledge, this was the first trial to assess a MedDiet in adults with RA residing in Ireland, where the Mediterranean dietary pattern is not very common. The study highlights that, although there is promise for the MedDiet in the management of RA, more studies are required to inform dietary recommendations and guidelines for a RA population. Moreover, our study demonstrated that nutrition counselling offered by a qualified RD intervention was successful at improving the dietary habits, therefore, implementation of dietary counselling should be considered and advocated for as a potentially important management approach in the treatment plan of RA.

### Conclusion

In conclusion, this study provides emerging data of the beneficial effects of optimising dietary quality under dietetic supervision in people with RA. Future studies considering objective outcomes such biochemical markers to confirm the results of this study and to assess physiological benefits of dietary prescription in this patient group. While results from the MEDRA study suggest that adopting a dietary pattern reflecting a healthy diet may be beneficial for RA, mechanisms by which improving dietary quality may achieve the observed improvements have yet to be fully investigated.

## Data Availability

All data produced in the present study are available upon reasonable request to the authors

## Footnotes

### Contributors

TR: conceptualisation, methodology, investigation, resources, data collection, data analysis, and writing (original draft, review and editing). AT: conceptualisation, methodology, investigation, resources and writing (review and editing), supervision and project administration, guarantor. AG, LL, AF, EG, NK: investigation and resources. All authors contributed to data interpretation and gave the final approval of the manuscript.

## References

1. Bax, M., et al., Genetics of rheumatoid arthritis: what have we learned? Immunogenetics, 2011. 63(8): p. 459–466.

2. de Reumatología, S.E., Eli Lilly And Company: RA Matters Survey Uncovers What Matters Most To Over 5,000 People Living With Rheumatoid Arthritis.

3. Fermoyle, V., Quality of Life in adults with Rheumatoid Arthritis: A closer Look at potential predictive factors. 2014.

4. Cherukumilli, V.S. and A. Kavanaugh, Elderly onset rheumatoid arthritis, in Geriatric Rheumatology. 2011, Springer. p. 145–150.

5. Sharif, K., et al., Rheumatoid arthritis in review: Clinical, anatomical, cellular and molecular points of view. Clinical Anatomy, 2018. 31(2): p. 216–223.

6. Lwin, M.N., et al., Rheumatoid arthritis: the impact of mental health on disease: a narrative review. Rheumatology and Therapy, 2020. 7(3): p. 457–471.

7. Ahlstrand, I., et al., Pain and daily activities in rheumatoid arthritis. Disability and Rehabilitation, 2012. 34(15): p. 1245–1253.

8. Semb, A.G., et al., Atherosclerotic cardiovascular disease prevention in rheumatoid arthritis. Nature Reviews Rheumatology, 2020. 16(7): p. 361–379.

9. Combe, B., Early rheumatoid arthritis: strategies for prevention and management. Best practice & research Clinical rheumatology, 2007. 21(1): p. 27–42.

10. Ferro, F., et al., One year in review 2017: novelties in the treatment of rheumatoid arthritis. Clin Exp Rheumatol, 2017. 35(5): p. 721–734.

11. Shams, S., et al., The therapeutic landscape of rheumatoid arthritis: current state and future directions. Frontiers in Pharmacology, 2021. 12: p. 1233.

12. Gwinnutt, J.M., et al., 2021 EULAR recommendations regarding lifestyle behaviours and work participation to prevent progression of rheumatic and musculoskeletal diseases. Annals of the rheumatic diseases, 2022.

13. Vlieland, T.P.V. and C.H. van den Ende, Nonpharmacological treatment of rheumatoid arthritis. Current opinion in rheumatology, 2011. 23(3): p. 259–264.

14. Tedeschi, S.K. and K.H. Costenbader, Is there a role for diet in the therapy of rheumatoid arthritis? Current rheumatology reports, 2016. 18(5): p. 1–9.

15. Vitetta, L., et al., Dietary recommendations for patients with rheumatoid arthritis: a review. Nutr Diet Suppl, 2012. 4(4): p. 1–15.

16. Philippou, E., et al., Rheumatoid arthritis and dietary interventions: systematic review of clinical trials. Nutrition reviews, 2021. 79(4): p. 410–428.

17. Raad, T., et al., Dietary interventions with or without omega-3 supplementation for the management of rheumatoid arthritis: A systematic review. Nutrients, 2021. 13(10): p. 3506.

18. Tsigalou, C., et al., Mediterranean diet as a tool to combat inflammation and chronic diseases. An overview. Biomedicines, 2020. 8(7): p. 201.

19. Rosato, V., et al., Mediterranean diet and cardiovascular disease: a systematic review and meta-analysis of observational studies. European journal of nutrition, 2019. 58(1): p. 173–191.

20. Daien, C., et al., Dietary recommendations of the French Society for Rheumatology for patients with chronic inflammatory rheumatic diseases. Joint bone spine, 2022. 89(2): p. 105319.

21. Schönenberger, K.A., et al., Effect of Anti-Inflammatory Diets on Pain in Rheumatoid Arthritis: A Systematic Review and Meta-Analysis. Nutrients, 2021. 13(12): p. 4221.

22. Forsyth, C., et al., The effects of the Mediterranean diet on rheumatoid arthritis prevention and treatment: a systematic review of human prospective studies. Rheumatology international, 2018. 38(5): p. 737–747.

23. Flynn, M.A., et al., Revision of food-based dietary guidelines for Ireland, Phase 2: recommendations for healthy eating and affordability. Public health nutrition, 2012. 15(3): p. 527–537.

24. McDougall, J., et al., Effects of a very low-fat, vegan diet in subjects with rheumatoid arthritis. The Journal of Alternative & Complementary Medicine, 2002. 8(1): p. 71–75.

25. Sköldstam, L., L. Hagfors, and G. Johansson, An experimental study of a Mediterranean diet intervention for patients with rheumatoid arthritis. Annals of the rheumatic diseases, 2003. 62(3): p. 208–214.

26. Michalsen, A., et al., Mediterranean diet or extended fasting’s influence on changing the intestinal microflora, immunoglobulin A secretion and clinical outcome in patients with rheumatoid arthritis and fibromyalgia: an observational study. BMC complementary and alternative medicine, 2005. 5: p. 1–9.

27. McKellar, G., et al., A pilot study of a Mediterranean-type diet intervention in female patients with rheumatoid arthritis living in areas of social deprivation in Glasgow. Annals of the rheumatic diseases, 2007. 66(9): p. 1239–1243.

28. Larkin, L., et al., The impact of COVID-19 on clinical research: the PIPPRA and MEDRA experience. HRB Open Research, 2021. 4(55): p. 55.

29. Cuschieri, S., The CONSORT statement. Saudi journal of anaesthesia, 2019. 13(Suppl 1): p. S27.

30. Raad, T., et al., A randomised controlled trial of a Mediterranean Dietary Intervention for Adults with Rheumatoid Arthritis (MEDRA): Study protocol. Contemporary Clinical Trials Communications, 2022: p. 100919.

31. Aletaha, D., et al., 2010 rheumatoid arthritis classification criteria: an American College of Rheumatology/European League Against Rheumatism collaborative initiative. Arthritis & rheumatism, 2010. 62(9): p. 2569–2581.

32. Kafatos, A., et al., Mediterranean diet of Crete: foods and nutrient content. Journal of the American Dietetic Association, 2000. 100(12): p. 1487–1493.

33. George, E., et al., A randomised controlled trial of a Mediterranean Dietary Intervention for Adults with Non Alcoholic Fatty Liver Disease (MEDINA): study protocol. 2022.

34. Henessay, M., 101+ Square meals evaluation Authors: Emma Hughes.

35. Casas, R., et al., Long-term immunomodulatory effects of a Mediterranean diet in adults at high risk of cardiovascular disease in the PREvención con DIeta MEDiterránea (PREDIMED) randomized controlled trial. The Journal of nutrition, 2016. 146(9): p. 1684–1693.

36. García-Conesa, M.-T., et al., Exploring the validity of the 14-item mediterranean diet adherence screener (Medas): A cross-national study in seven european countries around the mediterranean region. Nutrients, 2020. 12(10): p. 2960.

37. Allanore, Y., et al., Health Assessment Questionnaire-Disability Index (HAQ-DI) use in modelling disease progression in diffuse cutaneous systemic sclerosis: an analysis from the EUSTAR database. Arthritis research & therapy, 2020. 22(1): p. 1–11.

38. Tijhuis, G., et al., The validity of the rheumatoid arthritis quality of life (RAQoL) questionnaire. Rheumatology, 2001. 40(10): p. 1112–1119.

39. Dipietro, L., et al., A survey for assessing physical activity among older adults. Medicine & Science in Sports & Exercise, 1993.

40. Behrens, F., et al., Use of a “critical difference” statistical criterion improves the predictive utility of the Health Assessment Questionnaire-Disability Index score in patients with rheumatoid arthritis. BMC rheumatology, 2019. 3(1): p. 1–9.

41. Malaviya, A., et al., Prevalence of rheumatoid arthritis in the adult Indian population. Rheumatology international, 1993. 13(4): p. 131–134.

42. Casas, R., E. Sacanella, and R. Estruch, The immune protective effect of the Mediterranean diet against chronic low-grade inflammatory diseases. Endocrine, Metabolic & Immune Disorders-Drug Targets (Formerly Current Drug Targets-Immune, Endocrine & Metabolic Disorders), 2014. 14(4): p. 245–254.

43. Picchianti Diamanti, A., et al., Impact of Mediterranean diet on disease activity and gut microbiota composition of rheumatoid arthritis patients. Microorganisms, 2020. 8(12): p. 1989.

44. Farooqui, T. and A.A. Farooqui, Role of the Mediterranean Diet in the Brain and Neurodegenerative Diseases. 2017: Academic Press.

45. Mazzocchi, A., et al., The secrets of the Mediterranean diet. Does [only] olive oil matter? Nutrients, 2019. 11(12): p. 2941.

46. Matsumoto, Y., et al., Monounsaturated fatty acids might be key factors in the Mediterranean diet that suppress rheumatoid arthritis disease activity: The TOMORROW study. Clinical nutrition, 2018. 37(2): p. 675–680.

47. Kostoglou-Athanassiou, I., L. Athanassiou, and P. Athanassiou, The effect of omega-3 fatty acids on rheumatoid arthritis. Mediterranean journal of rheumatology, 2020. 31(2): p. 190.

48. Trichopoulou, A., et al., Definitions and potential health benefits of the Mediterranean diet: views from experts around the world. BMC medicine, 2014. 12(1): p. 1–16.

49. Bower, A., S. Marquez, and E.G. de Mejia, The health benefits of selected culinary herbs and spices found in the traditional Mediterranean diet. Critical reviews in food science and nutrition, 2016. 56(16): p. 2728–2746.

50. Gibney, M. and B. Sandström, A framework for food-based dietary guidelines in the European Union. Public Health Nutrition, 2001. 4(2a): p. 293–305.

51. Bruins, J., et al., The effects of lifestyle interventions on (long-term) weight management, cardiometabolic risk and depressive symptoms in people with psychotic disorders: a meta-analysis. PloS one, 2014. 9(12): p. e112276.

52. Sköldstam, L., et al., Weight reduction is not a major reason for improvement in rheumatoid arthritis from lacto-vegetarian, vegan or Mediterranean diets. Nutrition journal, 2005. 4(1): p. 1–6.

53. Tierney, M., A. Fraser, and N. Kennedy, Physical activity in rheumatoid arthritis: a systematic review. Journal of Physical Activity and Health, 2012. 9(7): p. 1036–1048.

54. Happell, B., C. Davies, and D. Scott, Health behaviour interventions to improve physical health in individuals diagnosed with a mental illness: A systematic review. International journal of mental health nursing, 2012. 21(3): p. 236–247.

55. Avery, L., et al., Changing physical activity behavior in type 2 diabetes: a systematic review and meta-analysis of behavioral interventions. Diabetes care, 2012. 35(12): p. 2681–2689.

56. Dale, H., L. Brassington, and K. King, The impact of healthy lifestyle interventions on mental health and wellbeing: a systematic review. Mental Health Review Journal, 2014.

57. García-Morales, J.M., et al., Effect of a dynamic exercise program in combination with Mediterranean diet on quality of life in women with rheumatoid arthritis. JCR: Journal of Clinical Rheumatology, 2020. 26(7S): p. S116–S122.

58. Clipper, B., The influence of the COVID-19 pandemic on technology: adoption in health care. Nurse Leader, 2020. 18(5): p. 500–503.

59. Hanlon, P., et al., Telehealth interventions to support self-management of long-term conditions: a systematic metareview of diabetes, heart failure, asthma, chronic obstructive pulmonary disease, and cancer. Journal of medical Internet research, 2017. 19(5): p. e6688.

60. Warner, M.M., et al., Reporting of telehealth-delivered dietary intervention trials in chronic disease: systematic review. Journal of medical Internet research, 2017. 19(12): p. e8193.

61. Kelly, J.T., et al., Dietitians Australia position statement on telehealth. Nutrition & Dietetics, 2020. 77(4): p. 406–415.

62. Chang, A.R., et al., Telehealth versus Self-Directed Lifestyle Intervention to Promote Healthy Blood Pressure: a Randomized Controlled Trial. medRxiv, 2022.

63. Poh, L., et al., An integrative review of experiences of patients with rheumatoid arthritis. International Nursing Review, 2015. 62(2): p. 231–247.

